# Self-learning on COVID-19 among medical students and their preparedness to participate in government’s COVID-19 response in Bhutan: a cross-sectional study

**DOI:** 10.1101/2020.09.07.20189936

**Authors:** Thinley Dorji, Saran Tenzin Tamang, TVSVGK Tilak

**Affiliations:** Department of Internal Medicine, Armed Forces Medical College, Maharashtra University of Medical Sciences, Pune, India; Kidu Mobile Medical Unit, His Majesty’s People’s Project, Thimphu, Bhutan; Faculty of Postgraduate Medicine, Khesar Gyalpo University of Medical Sciences of Bhutan, Thimphu, Bhutan

**Keywords:** medical education, medical school, online learning, social media, pandemic

## Abstract

**Background:** Bhutan lacks a medical school and all their medical students are trained outside in Sri Lanka, Bangladesh and India. When the COVID-19 pandemic let to closure of medical schools in these countries, the Bhutanese medical students were repatriated in March-April 2020. Upon return, they were kept in government-sponsored facility quarantine for 21 days. This study assessed their knowledge on COVID-19 as a part of self-learning and attitude as part of preparedness towards participation in government’s health response to COVID-19.

**Method:** This was a cross-sectional study among medical students who had returned to the country. This survey was conducted through an online questionnaire while the students were in 21-day facility quarantine. The sample size calculated was 129 and a convenient sampling was used. Knowledge was assessed using 20 questions, each scored 1/20. Cumulative score of score of ≥80% was categorized as “good knowledge”, score of ≥60 – 79% was considered “satisfactory knowledge”, and score < 60% was considered “poor knowledge.” Correlation between knowledge score and duration of clinical clerkship was tested using Pearson’s correlation coefficient. Attitude of students towards their willingness to participate in the national COVID-19 response were tested using rating scales. Data were analysed using Stata 13.1.

**Results:** 120 medical students responded to this survey (response rate = 93%). Eighty-eight (74%) had good knowledge, 28 (23%) had satisfactory knowledge and only four (3%) had poor knowledge on COVID-19. The students scored high on the symptomatology, mode of transmission, prevention and treatment options and on local epidemiology; and scored low on the forms of mechanical ventilations and on the home-management of non-critical cases. The knowledge score correlated with duration of clinical clerkship (r = 0.326, p = 0.001). The primary source of information were social media sites (102, 85%), television (94, 78%) and newspapers (76, 63%). The majority (78, 65%) were willing to participate in the government’s COVID-19 response but could not identify what roles they could play. The fear of contracting COVID-19 was reported in only in 8.7%.

**Conclusions:** The medical students had good knowledge on COVID-19 and were self-learned through social media, television and newspapers. The students held positive attitude towards participation in the government’s COVID-19 response.

## Introduction

The COVID-19 pandemic has brought an extraordinary situation across all walks of life and in the delivery of medical education [1]. In an effort to maintain social distancing and prevent community spread, countries have closed schools and colleges [2,3]. In South Asia, even before lockdown in India was announced in March 2020, schools and colleges including medical schools were closed. While some regulatory bodies have called for early graduation and registration of medical students to meaningful participation in the present pandemic, others have cautioned against taking up roles beyond their competence [2,4,5]. In Italy, at the height of health crisis when health system that was overwhelmed, medical students were fast tracked and recruited as doctors [6]. The urgency to recruit young health workers in the ongoing pandemic is also due to higher complication rates and mortality reported among elderly healthcare workers [5,6]. This has led to a tension between safeguarding education and responding to the demands on health service [7].

The pandemic situation has brought significant changes to medical education and particularly their transition from student to doctors [7,8]. With clinical rotations cancelled, students have reported reduced confidence and preparedness to work as doctors [7]. The online teaching-learning has been rapidly adopted across many colleges and institutions and has been particularly useful to those institutions with international students who returned to their native homes [9,10]. However, this is not served as a substitute for clinical exposure, clinical experience and collaborative learning opportunities [3]. The pandemic has delayed the graduation or licencing examinations of many students and this has a bearing on their subsequent applications for residency and postgraduate training programmes and the overall educational trajectory [7,11,12]. The disruption of medical education also has financial implications on the cost incurred for training and is a cause of anxiety in a significant proportion of students requiring mental health support [3,13].

Bhutan is a small country of population 0.79 million in the eastern Himalayas [14]. The Royal Government provides healthcare free of cost at all levels [15]. However, with a doctor-patient ratio of 5 per 10,000 population and with no medical colleges in the country, Bhutan still faces shortage of doctors and is dependent on the medical colleges in South Asia to train all of its doctors [15]. Presently, Bhutan has more than 180 medical students studying across Sri Lanka, Bangladesh and India [16].

After Bhutan reported its first case of COVID-19 in a tourist [17], the Royal Government took a decisive action to re-call forty-six postgraduate doctors studying outside the country [18] and more than 100 medical students studying in Sri Lanka, Bangladesh, Thailand and India [19]. Upon their return, like thousands of other Bhutanese who were evacuated to Bhutan, these doctors and medical students were kept in mandatory 21-day facility quarantine in hotels [20]. This study assessed the knowledge of medical students on COVID-19, the sources of information and their attitude towards participation in health response to COVID-19 in the country.

## Research method

### Study design

This was a cross-sectional study among Bhutanese medical students.

### Setting

#### General setting

Bhutan is a small country situated in the eastern Himalayas with a population of 0.7 million. Healthcare is provided free of cost at all levels, guided by the National Health Policy 2011 within the broader framework of overall national development and pursuit of happiness [21]. Healthcare is provided through a three-tiered system: primary health centres and 10-bedded hospitals at the primary healthcare level, district and general hospitals at secondary level, and referral hospitals with specialist services at the tertiary level. The three referral centres are located in geographically strategic locations in the west, east and the central regions. Specific COVID-19 management hospitals were designated across the country while the majority of the confirmed cases were treated at the Jigme Dorji Wangchuck National Referral Hospital, Thimphu.

#### Health human resources in Bhutan

Modern health system in Bhutan began in 1956 and there has been shortage of health human resources, particularly doctors [15,18,22]. All of Bhutan’s doctors are trained in other countries; all are employed by the Royal Civil Service Commission as the healthcare system is fully run by the government (no private practice is allowed). In 2020, there were 376 doctors in Bhutan with particular shortage of specialist doctors [23]. As of 2019, there are approximately 180 Bhutanese medical students in Sri Lanka, Bangladesh and India [16]. These students are fully or partially funded by the Royal Government while some are privately funded. The duration of undergraduate medical training is five years in Sri Lanka and four years in Bangladesh and India, followed by one year internship programme. The students can take up internship programme offered by the Faculty of Postgraduate Medicine, Khesar Gyalpo University of Medical Sciences of Bhutan at three tertiary hospitals in Bhutan.

As of March-April 2020, more than 100 medical students had returned to Bhutan [19]. As per the Royal Government of Bhutan’s COVID-19 response, these students were kept in government-sponsored mandatory facility quarantine in hotels in Paro and Thimphu dzongkhags for 21 days [20]. While in quarantine, persons returning from abroad were monitored daily for symptoms of COVID-19 and tested with RT-PCR if symptomatic and all were tested with rapid-antibody test at the end of quarantine period.

### Study site and study population

This study was conducted via online survey among medical students were in facility-quarantine in Paro and Thimphu *dzongkhags*.

### Sample size and sampling

For sample size calculation, we assumed that 50% of the respondents would have good knowledge on COVID-19. The sample size was calculated for proportions considering a 95% confidence level, 0.05 margin of error and finite correction for student population of 180 [16]. Allowing for a 5% dropout rate, the final sample size was 129. In the absence of the proper list of medical students coming into the country from various colleges and countries, convenience sampling was used.

### Study tool

The study questionnaire was designed to collect the respondent’s basic information, assessment of their knowledge and attitude and their source of information. The study tool was reviewed by seven national experts on COVID-19 for its content and construct validity and quality of multiple choice questions. As a result of this exercise, two items were revised and seven items were dropped. The S-CVI for the revised questionnaire was 0.96.

Knowledge is tested using twenty multiple choice questions. The knowledge was assessed under the following domains: disease definition, causative agent, symptomatology, mode of transmission and incubation period, clinical course of disease, diagnostic tests and centres in the country where these tests were available, case management and mortality rate, preventive measures, global and local epidemiology of COVID-19 and the national hotline number for COVID-19 related information and issues. We also assessed the common sources of COVID-19. Attitude was tested using rating scales against given statements on their willingness and fear to participate in COVID-19 responses, what role they would play and if medical students can make meaningful contributions in the pandemic settings in Bhutan. We also assessed if the students had received training on the use of personal protective equipment for COVID-19 care at their colleges.

### Data collection

Data were collected using Google Forms that were emailed to the students through their student representatives. Online data collection allowed social distancing at a time when quarantine facilities were out of bound for visitors.

### Data processing and analysis

The data were extracted from Google Forms and analysed in STATA Version 13.1 (StataCorp, Stata Statistical Software). Each knowledge question was scored 1/20. A cumulative score of ≥80% was categorized as “good knowledge”, score of ≥60 – 79% was considered “satisfactory knowledge”, and score < 60% was considered “poor knowledge.” Correlation between knowledge score and duration of clinical clerkship was tested using Pearson’s correlation coefficient. Results with *p*-value less than 0.05 were considered significant. Themes around attitude of students towards their willingness to participate in the national COVID-19 response are presented as frequencies and percentages.

### Ethics considerations

Ethics approval was obtained from the Research Ethics Board of Health, Ministry of Health, Bhutan. Informed consent was taken from the study participants as per the consent process approved by the ethics committee. For confidentiality, the data were anonymized and only the pooled/aggregate results are presented.

## Results

There were 120 medical students who responded to survey questionnaire (response rate = 93%). The mean age was 22 (±2) years; the proportion of students were distributed almost equally across first to fifth year of study; the median duration of clinical clerkship attended was 8 months (IQR 1, 18 months). The basic profile of the students and the country of study is shown in Table 1.

**Table 1.**
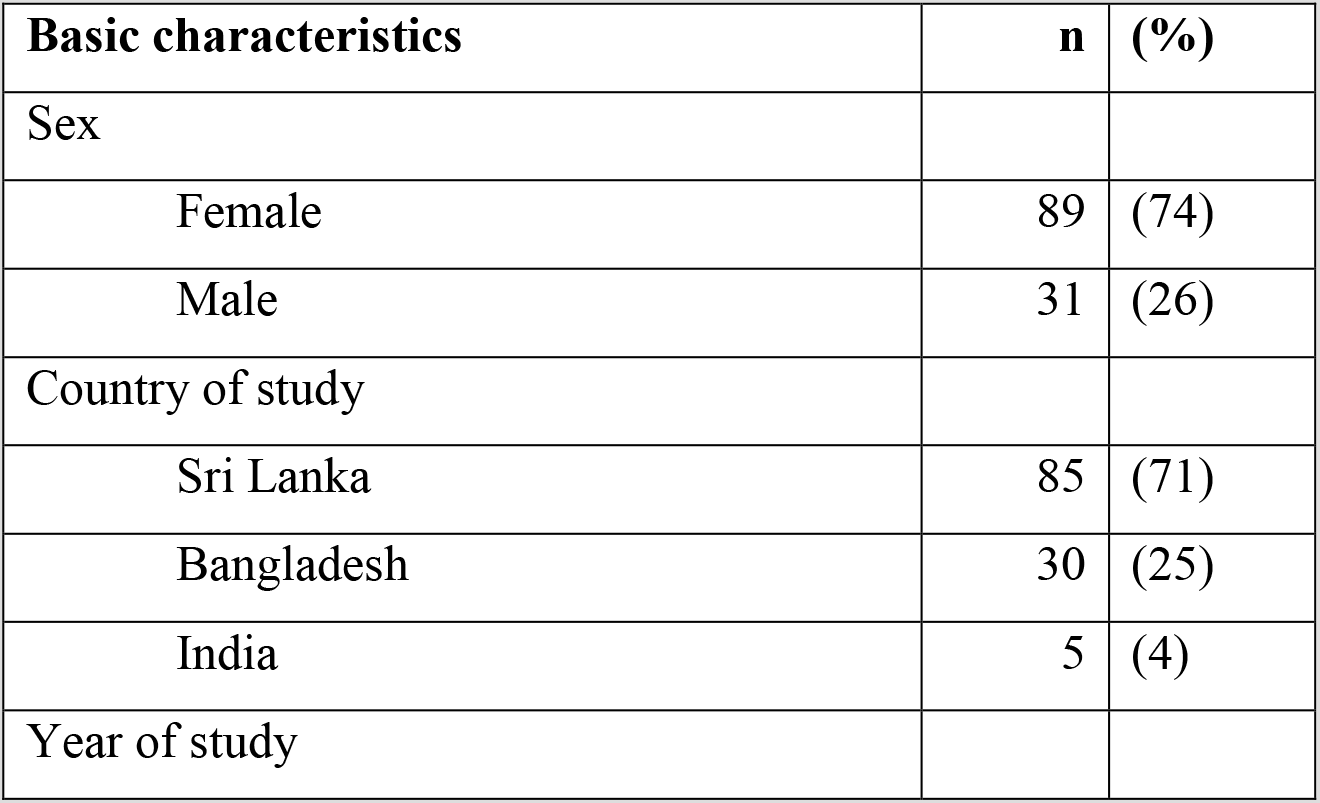
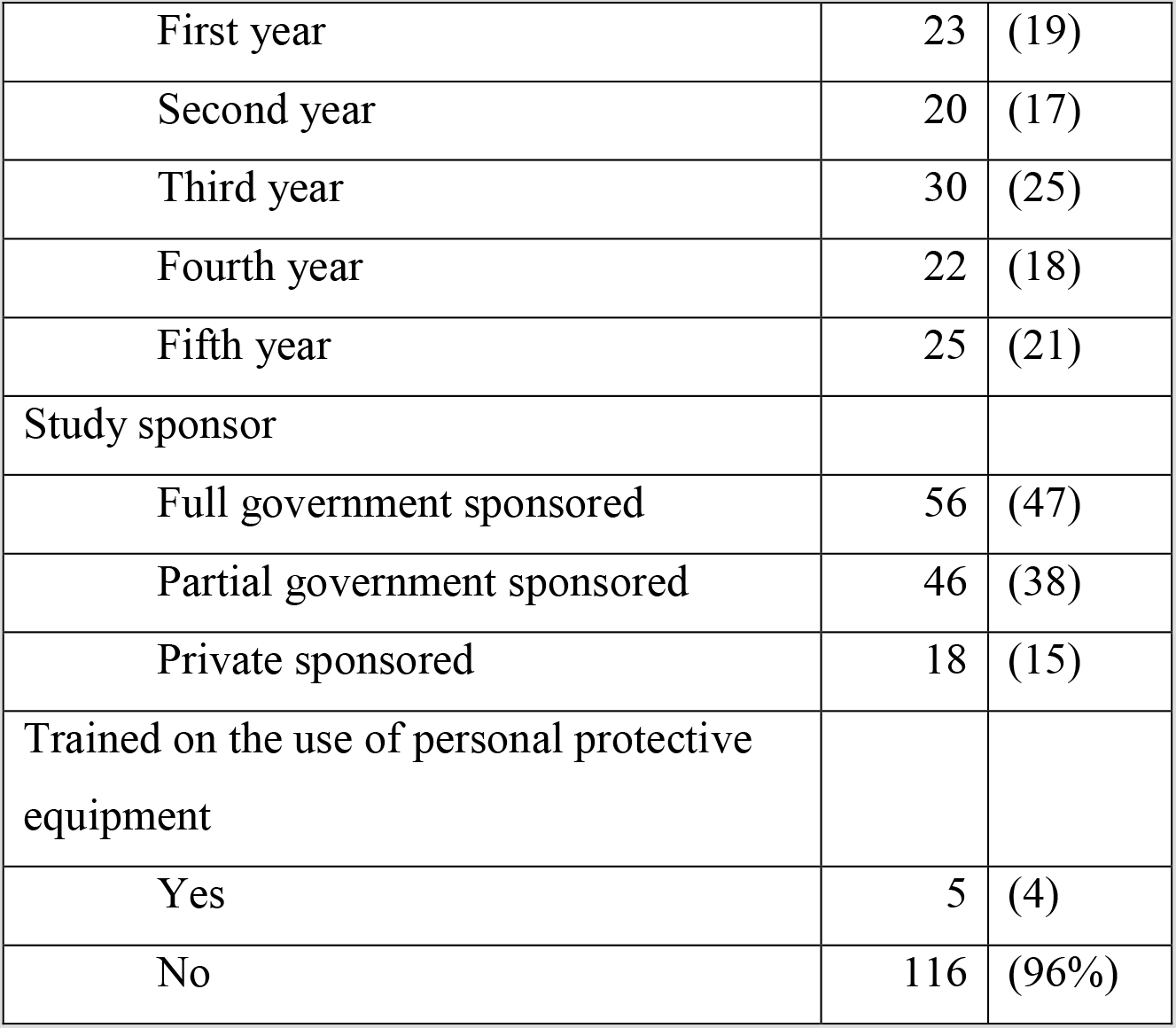
Basic profile of Bhutanese medical students studying Bachelor of Medicine and Bachelor of Surgery (MBBS) in Sri Lanka, Bangladesh and India surveyed for the COVID-19 KA study, April 2020

| Basic characteristics | n | (%) |
| --- | --- | --- |
| Sex |  |  |
| Female | 89 | (74) |
| Male | 31 | (26) |
| Country of study |  |  |
| Sri Lanka | 85 | (71) |
| Bangladesh | 30 | (25) |
| India | 5 | (4) |
| Year of study |  |  |
| First year | 23 | (19) |
| Second year | 20 | (17) |
| Third year | 30 | (25) |
| Fourth year | 22 | (18) |
| Fifth year | 25 | (21) |
| Study sponsor |  |  |
| Full government sponsored | 56 | (47) |
| Partial government sponsored | 46 | (38) |
| Private sponsored | 18 | (15) |
| Trained on the use of personal protective equipment |  |  |
| Yes | 5 | (4) |
| No | 116 | (96%) |

### Knowledge on COVID-19

Eighty-eight students (74%) had good knowledge, 28 (23%) had satisfactory knowledge and only four (3%) had poor knowledge on COVID-19. The students scored high on the symptomatology, mode of transmission, prevention and treatment options and on factors of local epidemiology such as all cases being imported, select centres with RT-PCR facilities and local COVID-19 hotlines. The students scored the lowest on the forms of mechanical ventilations and on the World Health Organization policy of home-management of non-critical COVID-19 cases. The knowledge score was significantly correlated with duration of clinical clerkship (r = 0.326, p = 0.001) and not related to sex, age or country of study. The details on knowledge score is shown in Table 2.

**Table 2.**
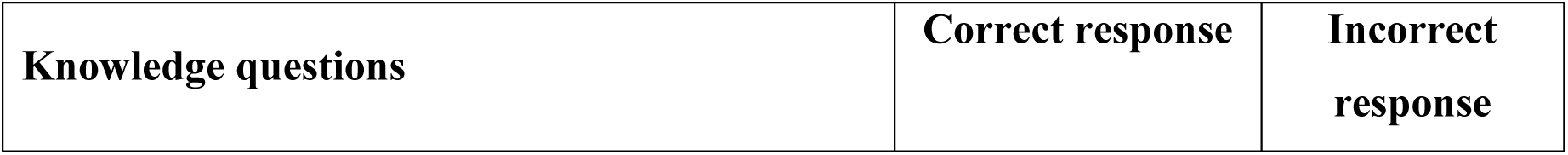
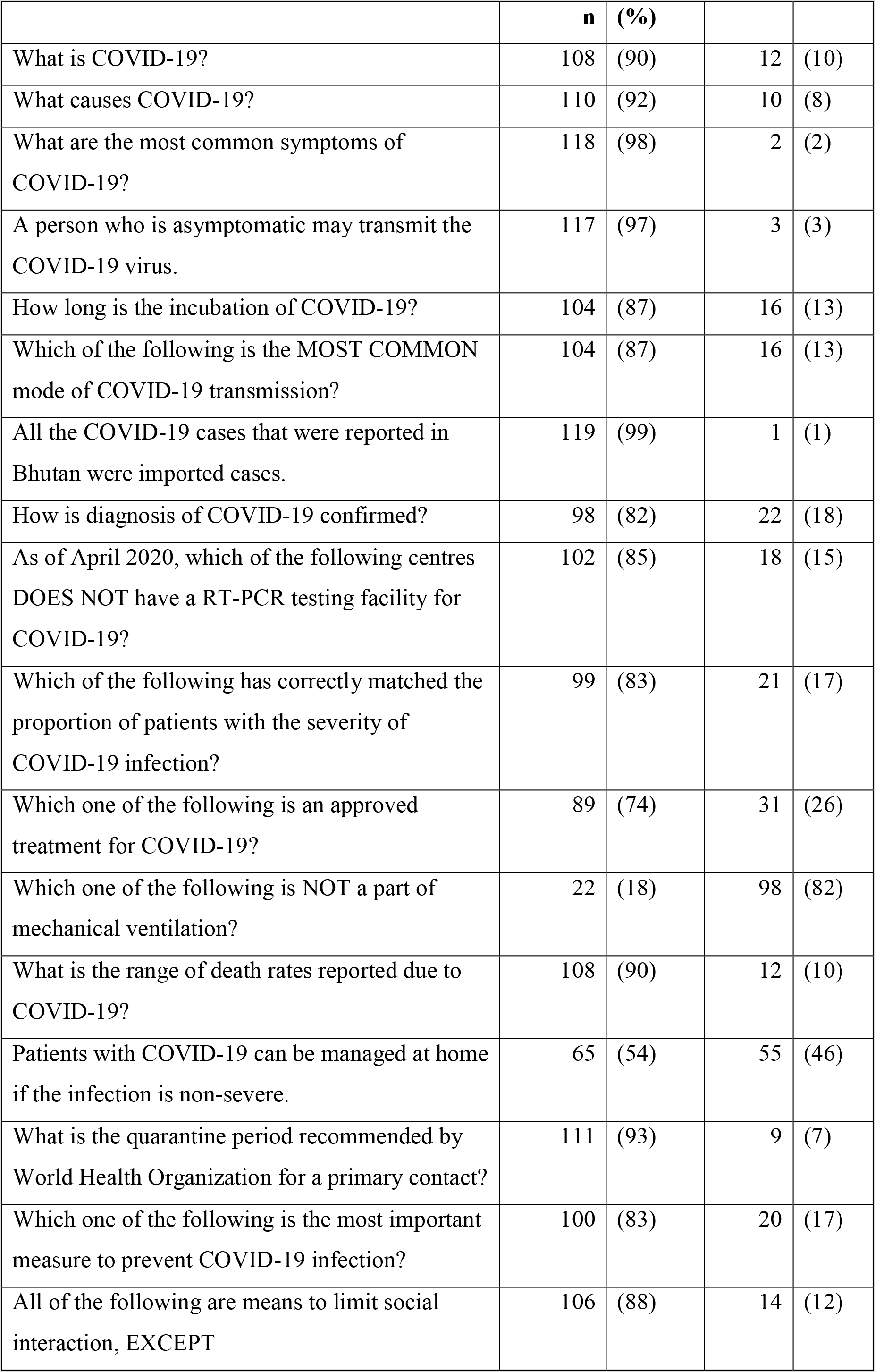
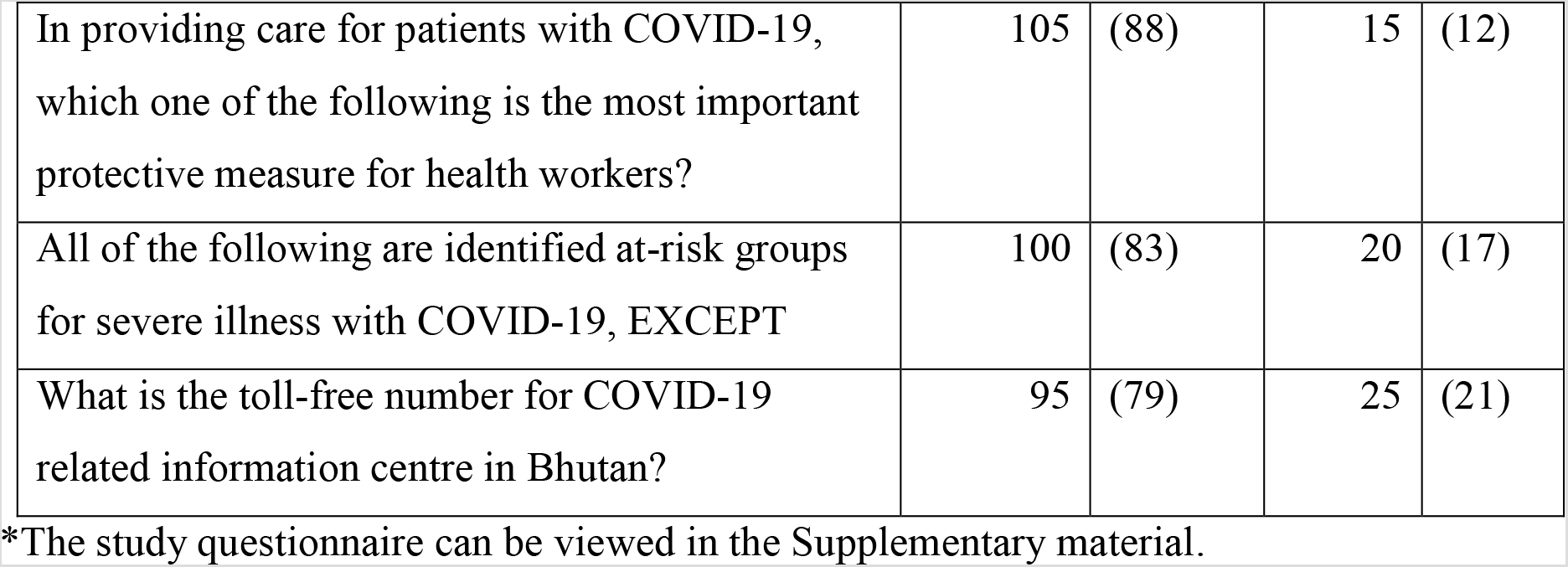
Assessment of knowledge on COVID-19 among Bhutanese medical students studying Bachelor of Medicine and Bachelor of Surgery (MBBS) in Sri Lanka, Bangladesh and India surveyed for the COVID-19 KA study, April 2020*

|  | **n** | **(%)** |  |  |
| --- | --- | --- | --- | --- |
| What is COVID-19? | 108 | (90) | 12 | (10) |
| What causes COVID-19? | 110 | (92) | 10 | (8) |
| What are the most common symptoms of COVID-19? | 118 | (98) | 2 | (2) |
| A person who is asymptomatic may transmit the COVID-19 virus. | 117 | (97) | 3 | (3) |
| How long is the incubation of COVID-19? | 104 | (87) | 16 | (13) |
| Which of the following is the MOST COMMON mode of COVID-19 transmission? | 104 | (87) | 16 | (13) |
| All the COVID-19 cases that were reported in Bhutan were imported cases. | 119 | (99) | 1 | (1) |
| How is diagnosis of COVID-19 confirmed? | 98 | (82) | 22 | (18) |
| As of April 2020, which of the following centres DOES NOT have a RT-PCR testing facility for COVID-19? | 102 | (85) | 18 | (15) |
| Which of the following has correctly matched the proportion of patients with the severity of COVID-19 infection? | 99 | (83) | 21 | (17) |
| Which one of the following is an approved treatment for COVID-19? | 89 | (74) | 31 | (26) |
| Which one of the following is NOT a part of mechanical ventilation? | 22 | (18) | 98 | (82) |
| What is the range of death rates reported due to COVID-19? | 108 | (90) | 12 | (10) |
| Patients with COVID-19 can be managed at home if the infection is non-severe. | 65 | (54) | 55 | (46) |
| What is the quarantine period recommended by World Health Organization for a primary contact? | 111 | (93) | 9 | (7) |
| Which one of the following is the most important measure to prevent COVID-19 infection? | 100 | (83) | 20 | (17) |
| All of the following are means to limit social interaction, EXCEPT | 106 | (88) | 14 | (12) |
| In providing care for patients with COVID-19, which one of the following is the most important protective measure for health workers? | 105 | (88) | 15 | (12) |
| All of the following are identified at-risk groups for severe illness with COVID-19, EXCEPT | 100 | (83) | 20 | (17) |
| What is the toll-free number for COVID-19 related information centre in Bhutan? | 95 | (79) | 25 | (21) |

\*The study questionnaire can be viewed in the Supplementary material.

The sources of information were social media sites – Facebook pages of the Ministry of Health (102, 85%) and the Prime Minister’s Office (80, 67%), television (94, 78%) and newspapers (76, 63%). Less than half of the students reported reading scientific journals for information (52, 43%). Only a quarter of the students (31, 26%) had attended lectures/symposia on COVID-19 at their colleges. The details of the sources of COVID-19 information are shown in Table 3.

**Table 3.**
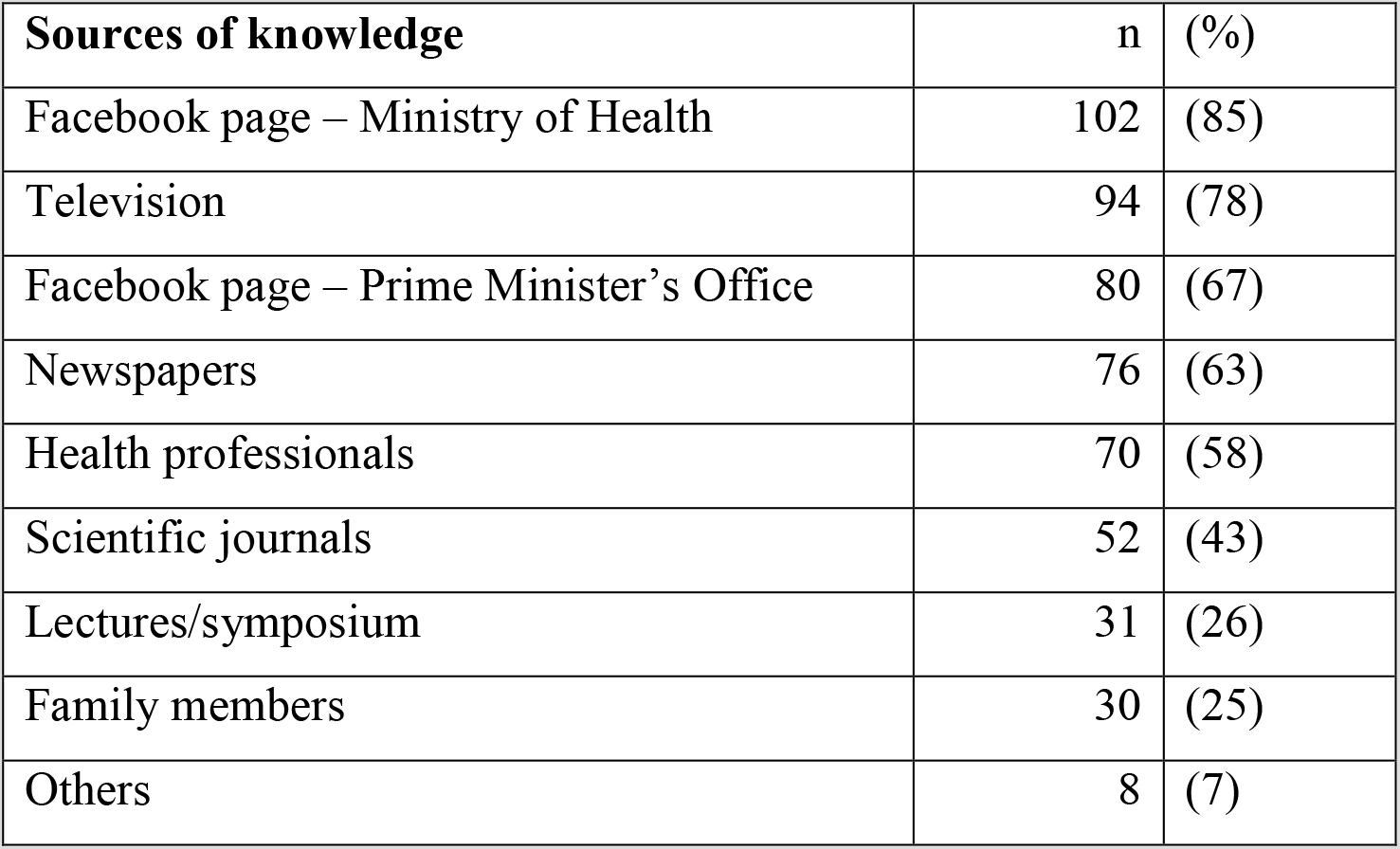
Sources of knowledge on COVID-19 reported by Bhutanese medical students studying Bachelor of Medicine and Bachelor of Surgery (MBBS) in Sri Lanka, Bangladesh and India surveyed for the COVID-19 KA study, April 2020

| Sources of knowledge | n | (%) |
| --- | --- | --- |
| Facebook page – Ministry of Health | 102 | (85) |
| Television | 94 | (78) |
| Facebook page – Prime Minister’s Office | 80 | (67) |
| Newspapers | 76 | (63) |
| Health professionals | 70 | (58) |
| Scientific journals | 52 | (43) |
| Lectures/symposium | 31 | (26) |
| Family members | 30 | (25) |
| Others | 8 | (7) |

### Attitude towards participation in COVID-19 response in the country

The majority (78, 65%) responded that they should participate in the government’s COVID-19 response and were willing to serve (92, 77%) in any part of the country. However, the students could not identify what roles they could play: clinical work (19, 16%) or advocacy and communications (33, 37%) but half of them felt that they may not make meaningful contributions in the COVID-19 response (57, 48%). The fear of contracting COVID-19 was reported in only a small proportion of students (8 7%). The details of the ratings on attitude statements are shown in Table 4.

**Table 4.** Attitude towards participation in COVID-19 response among Bhutanese medical students studying Bachelor of Medicine and Bachelor of Surgery (MBBS) in Sri Lanka, Bangladesh and India surveyed for the COVID-19 KA study, April 2020

| Attitude statements | Agree |  | Neutral |  | Disagree |  |
| --- | --- | --- | --- | --- | --- | --- |
|  | n | (%) | n | (%) | n | (%) |
| Medical students should participate in the COVID-19 response. | 78 | (65) | 41 | (34) | 1 | (1) |
| Medical students should be assigned only clinical work (taking care of patients) during the COVID-19 response effort. | 19 | (16) | 17 | (14) | 84 | (70) |
| Medical students should only work in communication and advocacy in the community on COVID-19 instead of the hospital. | 33 | (37) | 62 | (52) | 25 | (21) |
| I am willing to serve in any part (rural/urban) of the country wherever | 92 | (77) | 24 | (20) | 3 | (3) |
| required during the COVID-19 response effort. |  |  |  |  |  |  |
| In the current settings, medical students cannot make meaningful contributions in the COVID-19 response. | 16 | (13) | 47 | (39) | 57 | (48) |
| I am afraid that I might get COVID-19 if I visit the hospital. | 8 | (7) | 47 | (39) | 65 | (54) |

## Discussion

### Knowledge on COVID-19

The overall knowledge on COVID-19 was high amongst Bhutanese medical students studying in the three South Asian countries: Sri Lanka, Bangladesh and India. The students knew about the symptoms, mode of transmission, causative agent, confirmation of diagnosis and preventive measures. The students had good knowledge on the local epidemiology in Bhutan – cases being imported ones, the regional centres where RT-PCR facilities were available and the national COVID-19 hotline number. The knowledge was poor on options identifying the types of mechanical ventilation and that World Health Organization recommendation on home-based management of non-severe cases.

As the wave of the pandemic spread across South Asia, our study reflects the trend of medical learning among students from three countries when the pandemic disrupted their medical education. The learning pattern reported reflects a unique trend where social media information have contributed to more learning than conventional means such as lectures/symposia or the scientific literature. The majority of the students reported social media, television and newspapers as the source of information. This is similar to a survey involving 1404 medical students in Jordan and 1541 medical students in Turkey where the majority relied on online sources and social media platform for COVID-19 related information [11,24]. In Bhutan, the official Facebook page of the Ministry of Health and the Prime Minister’s Office carried information that were designed by their medical teams. This demonstrates the increasing usefulness of real-time sharing of information to students and knowledge transfer through social media [11,25]. The consumption of scientific journals (43%) was relatively higher than 27% reported among students in Jordan [24]. At the time of this study, most of the medical colleges had not initiated online lectures and only a quarter of the students had received information through their teachers.

Medical students are future healthcare providers and need comprehensive knowledge on both the clinical and public health management of emerging and re-emerging diseases. This study demonstrated that students had self-sufficiency and were able to self-learn adequate knowledge on a topic that was new and in a scenario that was hitherto not faced. Integration of learning with online tools goes beyond traditional roles of students [25,26] and is an important attribute adaptation to becoming successful doctors for the 21^st^ century [11,27]. However, as the World Health Organization has cautioned, medical students must learn to assess the reliability of the source of information at times of infodemic with many unregulated contents available online [28].

### Attitude towards engagement in COVID-19 activities

The students held positive attitudes towards participation in the national COVID-19 response. This is similar to findings from a survey of 179 medical students in Singapore where two-thirds reported that it was their professional responsibility to be a part of the medical team and participate in clinical activities in hospitals [29]. It has been suggested that in places where there are acute shortages, the medical students may be tasked with care of routine patients to free the other health workers for COVID-19 management [5,6]. In our study, however, the students could not identify whether they should shoulder clinical job responsibilities or take on communications and advocacy work and only 13% believed that they can make meaningful contributions. In a study in Turkey, a quarter of the students (24.3%) reported that they are not competent by any means to handle clinical responsibilities where as others reported that they would be happy to work in the emergency department [11]. Therefore, engagement of medical students in healthcare settings advisable only after thorough assessment of student’s readiness and the severity of needs and shortages in the system. It is suggested that medical students can assist with routine outpatient clinical care, care of chronic patients and routine antenatal checks of pregnant women. Many of these task can be performed via telemedicine and involves no risk of infectious transmission [26]. Advanced medical students may be engaged in inpatient care [26].

One of the reasons that may deter medical students from returning to clinical settings is their fear of contracting COVID-19. Among medical students, fear was reported in 1.3% medical students in Turkey and anxiety in 24.9% of medical students in a study in China [11,13]. In our study, 7% of students reported fear of contracting COVID-19. In our survey, fear was assessed against a single rating statement while instruments such as FCoV-19S, validated among medical students in Vietnam was not available at the time of data collection [30].

### Preparedness of medical students in clinical engagement

The involvement of medical students in clinical roles in the COVID-19 pandemic varies across countries and their regulatory bodies. Many have argued that medical students are not certified practitioners, increases the risk of exposure and consumption of PPEs and might act as additional vectors for viral transmission [11,26]. The proportion of medical students who received training on the use of personal protective equipment was very low: 4% in our study, and 7% among final year students in Turkey [11]. If the government were to engage medical students in clinical settings, proper inductions, training on training on the infection control measures and the use of PPEs and clear guidance on working within their competencies are required [7,31]. It is suggested that medical students be given targeted volunteering opportunities that supplement their educational needs and specific learning objectives [7]. Experience from Denmark shows that after adequate training on infection control and emphasis on the role of medical students in the pandemic emergency healthcare workforce, two thirds of students had volunteered to work in pandemic emergency departments [31].

### Impact of COVID-19 on medical education of Bhutanese students

As the wave of this pandemic spread across the globe, colleges and universities have implemented various strategies to provide medical education. While some medical colleges have engaged their students through online teaching platforms [32], some have been left without any engagement. While some are concerned over loss of academic year and would want to return to their colleges, some remain reluctant. In an assessment of student’s preference to return to clinical work in Singapore, one third of medical students perceived high personal risk of infection and preferred not to return to clinical education settings [29].

For Bhutan, the disruption of undergraduate medical education has major impact on the supply of doctors. Bhutan is dependent on the yearly supply of these medical graduates to address to the need of the country and the expanding medical services. With these medical students likely to take additional time to graduate, it would entail additional cost in training them to obtain the MBBS degree. This would delay their Royal Civil Service examinations and their entry into service and subsequently residency training programme in later years. Such impacts and disruption in the medical education and certification of doctors have reported an increase in anxiety and mental health issues among medical students during the pandemic period [3,8].

One option for these students in Bhutan is to allow clinical clerkships in the teaching hospitals in Bhutan, as were done during events of disruptions of clinical clerkships in the US [8]. They may be accommodated in the three teaching hospitals in the country: Jigme Dorji Wangchuck National Referral Hospital, Thimphu; Central Regional Referral Hospital, Gelegphu; and Eastern Regional Referral Hospital, Monggar. However, with these hospitals are designed as teaching hospitals for only postgraduate students and are currently engaged in activities as they serve as COVID-19 focal hospitals. As a result, many of the students have remained unengaged in clinical settings.

The scenario of medical education has rapidly evolved with adoption of online teaching-learning tools as a means of social distancing [9] while several colleges have adopted and approved online assessment [7,32]. In the wake of national lockdowns and ban on international travel, the Sri Lanka Medical Council approved the award of MBBS degrees of students who gave their theory and clinical examination conducted online at the Khesar Gyalpo University of Medical Sciences of Bhutan, Thimphu and assessed by their teachers in respective colleges.

### Study limitations

As the scenario of COVID-19 situation progressed in South Asia and in Bhutan, the perception of these medical students might have changed over time. This paper was an attempt to understand their knowledge and attitude towards their participation in COVID-19 response in the context of Bhutan.

## Conclusion

The medical students had good knowledge on COVID-19 and the source of information were mostly social media online content. The students had positive attitude towards participating in the COVID-19 national response but did not identify any specific roles in which they can make meaningful contributions.

## Data Availability

The data used for this study is available as supplementary file (dta).

## Data availability statement

The data used for this study is available as supplementary file (dta).

## Declaration of interest

The authors declare that there are no competing interests.

## Financial/material support

There is no financial or material support for this study.

## Acknowledgements

The authors are grateful to the medical students who responded to the survey.

## Authors’ contributions

TD, STT and TVSVGKT were involved in the conception and design of this study and collection of data. TD analysed and interpreted the data and drafted the manuscript. All authors were involved in critically reviewing the paper. All authors read and approved the final manuscript.

## Notes

### Competing Interest Statement

The authors have declared no competing interest.

### Author Declarations

Approved by Research Ethics Board of Health, Ministry of Health, Royal Government of Bhutan, Thimphu

